# Survival prediction for bladder cancer using machine learning: development of BlaCaSurv online survival prediction application

**DOI:** 10.1101/2020.11.13.20231191

**Authors:** Ashis Kumar Das, Shiba Mishra, Devi Kalyan Mishra, Saji Saraswathy Gopalan

## Abstract

**Background:** Bladder cancer is the most common cancer of the urinary system among the American population and it is the fourth most common cause of cancer morbidity and the eight most common cause of cancer mortality among men. Using machine learning algorithms, we predict the five-year survival among bladder cancer patients and deploy the best performing algorithm as a web application for survival prediction.

**Methods:** Microscopically confirmed adult bladder cancer patients were included from the Surveillance Epidemiology and End Results (SEER) database (2000-2017) and randomly split into training and test datasets (70/30 ratio). Five machine learning algorithms (logistic regression, support vector machine, gradient boosting, random forest, and K nearest neighbor) were trained on features to predict five-year survival. The algorithms were compared with performance metrics and the best performing algorithm was deployed as a web application.

**Results:** A total of 52,529 patients were included in our study. The gradient boosting algorithm was the best performer in terms of predictive ability and discrimination. It was deployed as the survival prediction web application named BlaCaSurv (https://blacasurv.herokuapp.com/).

**Conclusions:** We tested several machine learning algorithms and developed a web application for predicting five-year survival for bladder cancer patients. This application can be used as a supplementary prognostic tool to clinical decision making.

## 1. Introduction

Bladder cancer is the most common cancer of the urinary system among the American population, and an estimated 81,400 new cases of bladder cancer and 17,980 deaths are expected to occur in the United States in 2020.^1^ It is the fourth most common cause of cancer morbidity and the eight most common cause of cancer mortality among men. The 5-year relative survival rate for bladder cancer is 77% and has remained at similar levels over the past two decades.^2^

It is essential to understand the prognostic factors for bladder cancer outcomes for risk stratification, effective planning of treatment and rehabilitation modalities. While there have been several studies translating the prognostic factors to predictive models on bladder cancer, very few have used machine learning, and none have developed an open access web application.^3,4^ Moreover, most of the published studies have used data from single centers using smaller sample sizes that may not be generalizable.^5^

Machine learning is an emerging technique to predict diseases, risk factors, drug response, patient survival and cost-effectiveness of interventions using hospital and medical databases.^6–10^ As an artificial intelligence tool, machine learning algorithms apply a predictive model to unseen data (test data), while learning from a set of data (training data).^11^ With respect to cancer research a growing body of literature has shown application of machine learning for predicting cancer survival from hospital records and registries.^12–17^ The largest publicly available source of cancer statistics in the United States is the Surveillance Epidemiology and End Result (SEER) with a representation of 28% of the population.^18^ Using SEER database, a few studies have applied machine learning to predict patient survival on various cancers.^14–16^ However, its application to bladder cancer has been limited.^5^ Using machine learning algorithms, we predicted the five-year survival among bladder cancer patients. In addition, we deployed the best performing algorithm as a web application for future validation and clinical use.

## 2. Methods

### 2.1. Patients

Patients for this study were selected from the Surveillance Epidemiology and End Result (SEER) 21 databases available for cases from 2000 through 2017.^19^ The case completeness rate for the SEER database is 98% and all patients were followed up for 10 years after routine treatment until death or loss to follow-up.^20^ These databases include patient details such as demographic background, tumor features, and survival.

Inclusion criteria for this study were microscopically confirmed bladder cancer patients aged 18 and above, and availability of the following information – age, sex, race, histologic type, tumor site, grade, tumor size, number of in-situ tumors, summary stage, type of surgery performed and survival months. Patients with incomplete or missing information based on the above criteria were excluded. In terms of the timeframe, we considered patients diagnosed between 2004 and 2012 so as to have adequate follow up period (5 years or more) after the diagnosis. After excluding patients that did not meet our inclusion criteria, 52,529 bladder cancer patients were included in our study.

### 2.2. Outcome variable

The outcome variable in our study was survival of five or more years among bladder cancer patients. Survival is a continuous variable in the SEER database with units given in months. We created a binary survival variable where any patient that had a survival of 60 or more months was coded “yes” or “no” otherwise.

### 2.3 Predictors

We included patient level demographic and tumor specific variables as predictors. Demographic predictors were sex, age (years at diagnosis), and race. Age was a continuous variable from 18 years up to 84 year, and then ages 85 and above were coded as 85+ in SEER dataset. There were six races – “Hispanic”, “non-Hispanic American Indian/Alaska native”, “non-Hispanic Asian or Pacific Islander”, “non-Hispanic black”, “non-Hispanic white” and “non-Hispanic unknown”.

Tumor specific variables that were available in the database were histologic type, site, grade, tumor size, number of in-situ tumors, summary stage and type of surgery performed. There were three histologic types – transitional cell carcinoma, squamous cell carcinoma and others. Possible sites of tumor were ten – trigone, dome, lateral wall, anterior wall, posterior wall, bladder neck, ureteric orifice, urachus, overlapping lesion of bladder and bladder NOS. Tumor grade consisted of five options – I, II, III, IV and unknown. There were seven tumor sizes – none, up to 10 mm, 11-20 mm, 21-30 mm, 31-40 mm, 41-50 mm and above 50 mm. Number of in-situ tumors could be either solitary or multiple. There were five summary stages as follows in situ, localized, regional, distant and unknown. Finally, three were six categories for the type of surgery performed – no surgery, TURBT, partial cystectomy, radical cystectomy, pelvic exenteration and other.

### 2.4 Statistical Methods

#### 2.4.1 Descriptive Analysis

We performed descriptive analyses of the predictors stratified by their groups. The correlation was tested among all predictors with Pearson’s correlation coefficient.

#### 2.4.2 Predictive Analysis

To predict the determinants of five-year survival for bladder cancer, we applied five commonly used supervised machine learning algorithms in cancer survival research – logistic regression, support vector machine (SVM), K neighbor classification (KNN), random forest, and gradient boosting. The choice of the algorithms was driven by lower training time as well as lower lag time when set up as an online application. For each algorithm, we performed hyperparameter tuning to find the best-fitting parameters.

Logistic regression is used for binary or categorical outputs. The algorithm fits the best model to describe the relationship between the output (outcome) and input (predictor) variables.^21^ The best fit parameters in our model were L2 regularization and a penalty strength of 5. Support vector machine (SVM) uses a hyperplane to separate the two classes of the output.^17^ The algorithm tries to maximize the distance between the hyperplane and the two closest data points from each class. The three important parameters in SVM are kernel (transforms data from a linear to a spatial form such as linear, radial, sigmoid, or polynomial), penalty (an error term) and gamma (a measure of model fitting). The best parameters in our SVM algorithm were radial kernel, penalty of 1 and gamma of 0.01. The majority class among its neighbors in KNN algorithm decides the class of a new observation.^22^ The three critical parameters for KNN are number of nearest neighbors (number of data points a new observation is associated with), distance metric (distance between the new observation and the nearest neighbors, e.g. Euclidean, Manhattan and Minkowski) and weights (contribution of the members in the neighborhood). The best parameters in our model were eight nearest neighbors, Euclidean metric and distance weights. Both random forest and gradient boosting use decision trees that are generated by recursively splitting the predictors.^23,24^ Learning dynamically from its precursor, each decision tree passes on the function to the following tree in gradient boosting. In random forest, the trees are created in parallel. The learning can be improved by adjusting the learning rate, number of trees, depth of each tree, and splitting sample. The final predictions are taken out of a weighted combination of these trees. While random forest combines the weights at the end of the process, gradient boosting on the other hand combines the weights along the way from the beginning. The best fit parameters for random forest were 80 decision trees, maximum depth of 25 for each tree, minimum leaf samples of five to split, minimum leaf sample of one and 0.15 learning rate. For the gradient boosting algorithm, the best fit parameters were 800 decision trees, maximum depth of eight for each tree, minimum leaf samples of three to split, four maximum features and 0.15 learning rate.

##### 2.4.2.5 Evaluation of the performance of the algorithms

We split the data into training (70 percent) and test cohorts (30 percent). Initially, the algorithms were trained on the training cohort and then were validated on the test cohort (new data) for determining predictions. For training, the data was passed through a five-fold cross validation where the data was split randomly into training and validation cohorts at 80/20 ratio five times. The final prediction came out of the cross-validated estimate. To deal with imbalanced data (27.8% patients did not survive five or more years against 72.2% that did), we applied two oversampling techniques called adaptive synthetic (ADASYN) method and synthetic minority oversampling technique (SMOTE) to enhance the learning on the training data.^25,26^ ADASYN creates synthetic samples adaptively based on the samples’ complexity in learning.^27^ SMOTE generates synthetic samples from the minority class (patients that did not survive five or more years in our data) considering the feature space similarities between nearest neighbors.^25^

We evaluated the performance of the algorithms using four metrics – area under receiver operator characteristic curve (AUC), accuracy, sensitivity and specificity. The receiver operator characteristic (ROC) curve plots true positivity on the vertical axis (Y-axis) versus false positivity on the horizontal axis (X-axis).^28^ AUC measures the area under the ROC curve that ranges from 0.5 to 1.0 with higher values meaning better discriminating ability of the algorithm. Accuracy is a measure of correct classification of non-survived cases as non-survived and survived cases as survived by the algorithm.^28^ Sensitivity depicts correct prediction of non-survived cases among all those who did not survive, whereas specificity demonstrates correct prediction of survival among all those who survived.

The statistical analyses were performed using Stata Version 15 (StataCorp LLC. College Station, TX), Python programming language Version 3.7.1 (Python Software Foundation, Wilmington, DE, USA). The web application was built using the Flask application for Python and deployed with Heroku server.

## 3. Results

In this section, we present the profile of patients, performance of the algorithms and our online bladder cancer survival prediction application.

### 3.1 Patient profile

Table 1 presents the demographic profile of the patients. The mean age of the patients was 68.6 years with a standard deviation of 11.6. Slightly below three-fourths were males (73.9%). Non-Hispanic Whites were the majority (83.7%) race followed by Hispanics (5.9%) and non-Hispanic Blacks (5.2%). Out of all histologic types, patients with transitional cell carcinoma were the majority group (94.5%). Apart from the not otherwise specified (NOS) site (29.3%), the most common site of tumor was the lateral wall (25.4%). Grade IV was the majority category (29.6%) followed by grades II (25.2%), III (20.4%), unknown (13.4%) and I (11.4%). Slightly below a fourth of the tumors were 21-30 mm in size (24%) and most were solitary in-situ tumors (63.2%). As far as the summary stage is concerned, the majority were in situ (47.8%), followed by localized (35.5%) and regional (11.8%). In terms of surgical treatment, the most (64.8%) underwent trans urethral resection of bladder tumor (TURBT). Closer to 72% of the patients in our sample had a survival of five or more years. The correlation coefficients between the predictors ranged from -0.06 to 0.2.

**Table 1.**
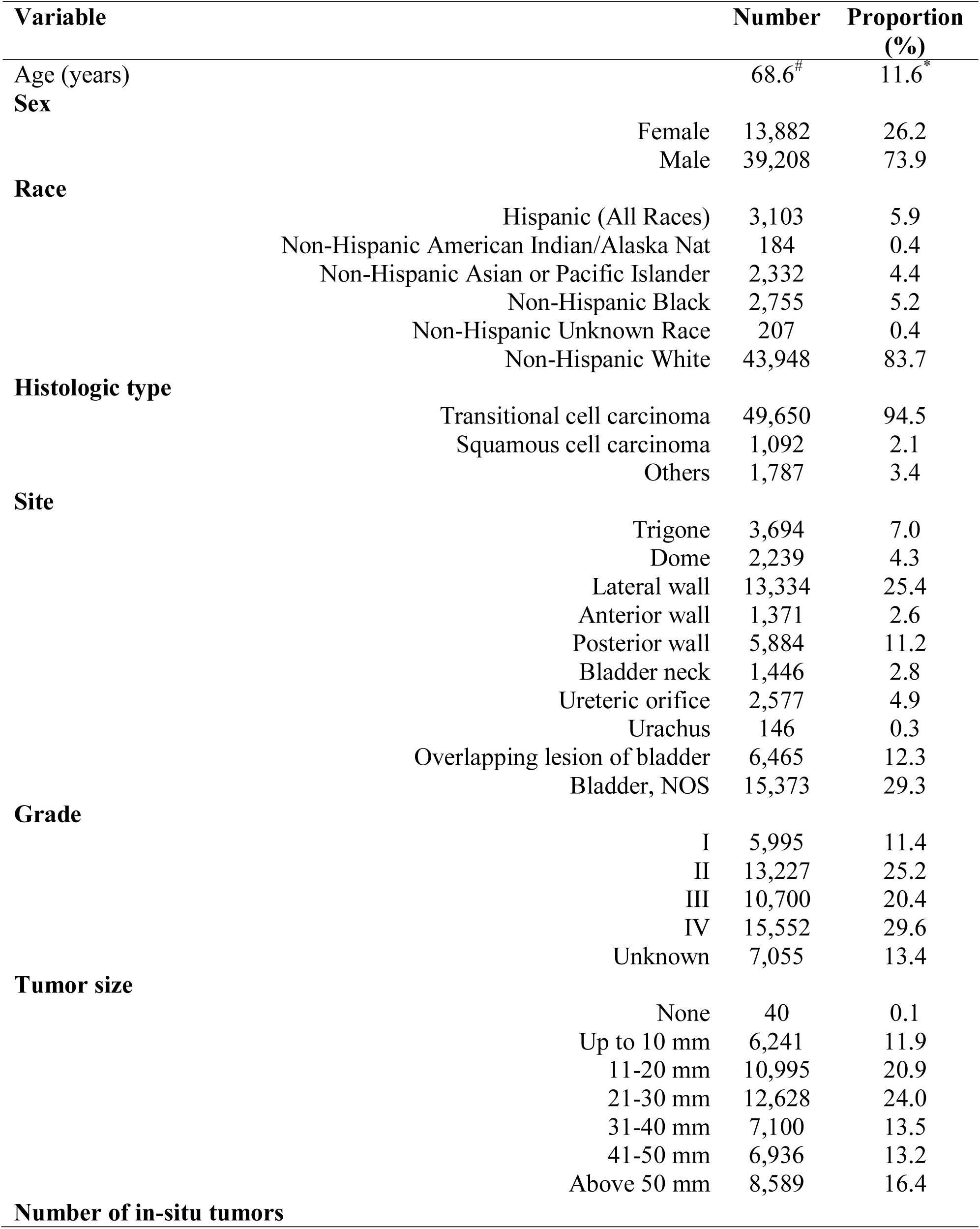
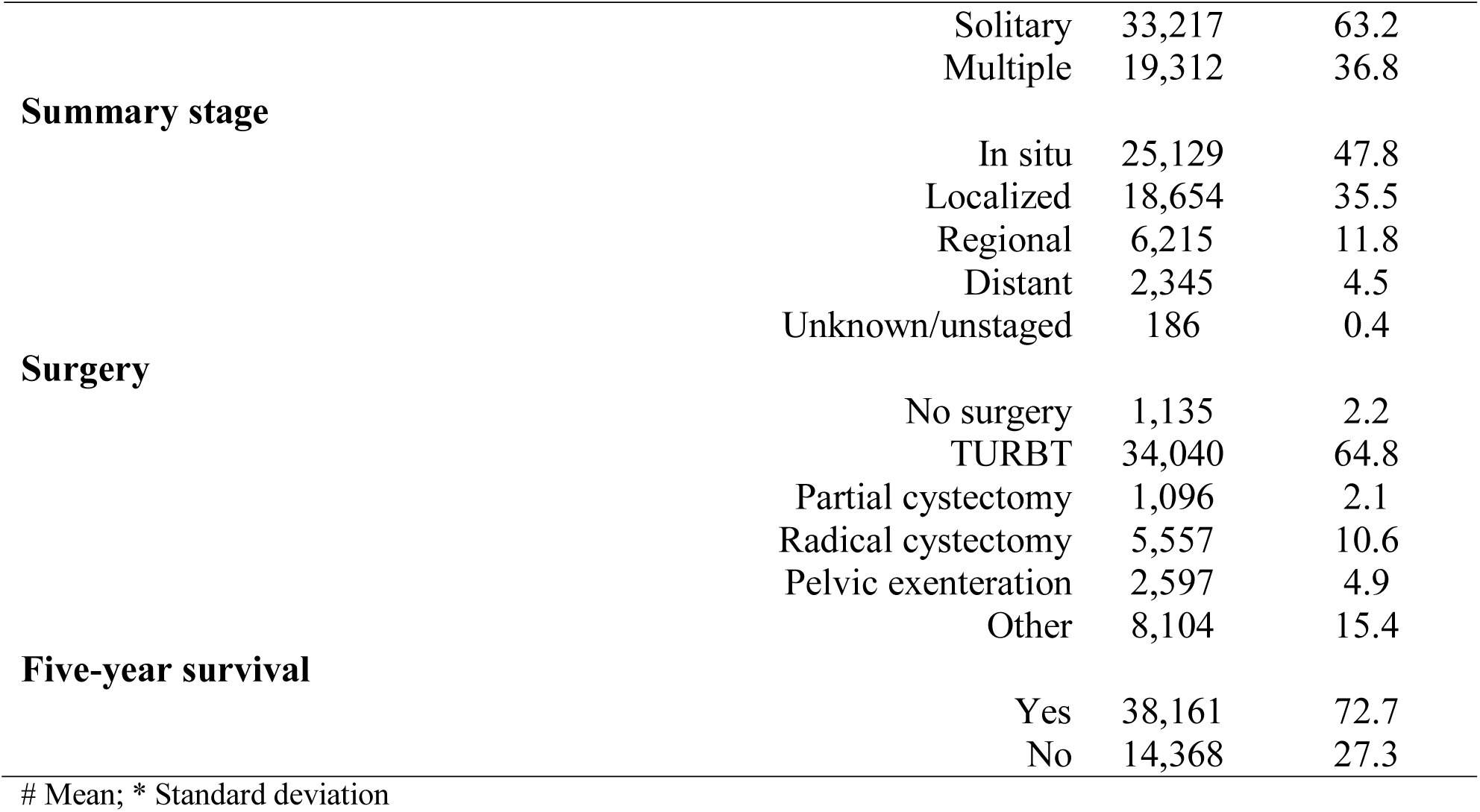
Patient profile.

### 3.2 Performance of the algorithms

Table 2 shows the performance metrics of the algorithms (logistic regression, support vector machine, K nearest neighbor, random forest and gradient boosting). The area under receiver operating characteristic curve (AUC) ranged from 0.859 to 0.924 with the best score for the gradient boosting (SMOTE) algorithm. Gradient boosting (SMOTE) algorithm also performed the best on accuracy (0.859) and sensitivity (0.863). Considering all the performance metrics, gradient boosting (SMOTE) was the best performing algorithm.

**Table 2.** Performance of the machine learning algorithms.

| Algorithm | Oversampling method | Area under ROC curve | Accuracy | Sensitivity | Specificity |
| --- | --- | --- | --- | --- | --- |
| **Logistic regression** | SMOTE<sup>#</sup> | 0.918 | 0.842 | 0.853 | 0.686 |
|  | ADASYN<sup>*</sup> | 0.907 | 0.828 | 0.847 | 0.691 |
| **Support vector machine** | SMOTE | 0.905 | 0.830 | 0.827 | 0.719 |
|  | ADASYN | 0.894 | 0.821 | 0.825 | 0.708 |
| **K nearest neighbor** | SMOTE | 0.875 | 0.812 | 0.790 | 0.638 |
|  | ADASYN | 0.859 | 0.800 | 0.767 | 0.651 |
| **Random forest** | SMOTE | 0.914 | 0.836 | 0.827 | 0.695 |
|  | ADASYN | 0.898 | 0.825 | 0.797 | 0.678 |
| **Gradient boosting** | SMOTE | 0.927 | 0.859 | 0.863 | 0.610 |
|  | ADASYN | 0.912 | 0.835 | 0.818 | 0.646 |

### 3.3 Online survival prediction application – BlaCaSurv

The best performing model, gradient boosting (SMOTE) was deployed as the online survival prediction application named as “**Bla**dder **Ca**ncer **Surv**ival - “BlaCaSurv” (https://blacasurv.herokuapp.com/). Figure 1 shows the user interface (UI). The UI has ten boxes for each input feature as drop-down menus. The features are age (continuous feature from 18 through 85), sex (two options – male and female), race (six options – Hispanic, non-Hispanic American Indian/Alaska native, non-Hispanic Asian or Pacific Islander, non-Hispanic Black, non-Hispanic white and unknown), histologic type (three options – transitional cell carcinoma, squamous cell carcinoma and others), site (ten options – trigone, dome, lateral wall, anterior wall, posterior wall, bladder neck, ureteric orifice, urachus, overlapping lesion of bladder and bladder NOS), grade (five options – I through IV and unknown), tumor size (seven options – none, up to 10 mm, 11-20 mm, 21-30 mm, 31-40 mm, 41-50 mm and above 50 mm), number of in-situ tumors (two options – solitary and multiple), summary stage (five options – in situ, localized, regional, distant and unknown/unstaged), and surgery performed (six options – no surgery, TURBT, partial cystectomy, radical cystectomy, pelvic exenteration and other). After selecting one option from each of the feature boxes and clicking the submit button, the application shows the estimated five-year survival probability in percentages. For instance, the application gives a five-year survival prediction of 40.88 % for a 60-year old male non-Hispanic Black patient with a grade 3 and regional tumor at the trigone. The tumor has multiple in-situ malignant lesions, shows transitional cell carcinoma in histology and TURBT as the surgical treatment.

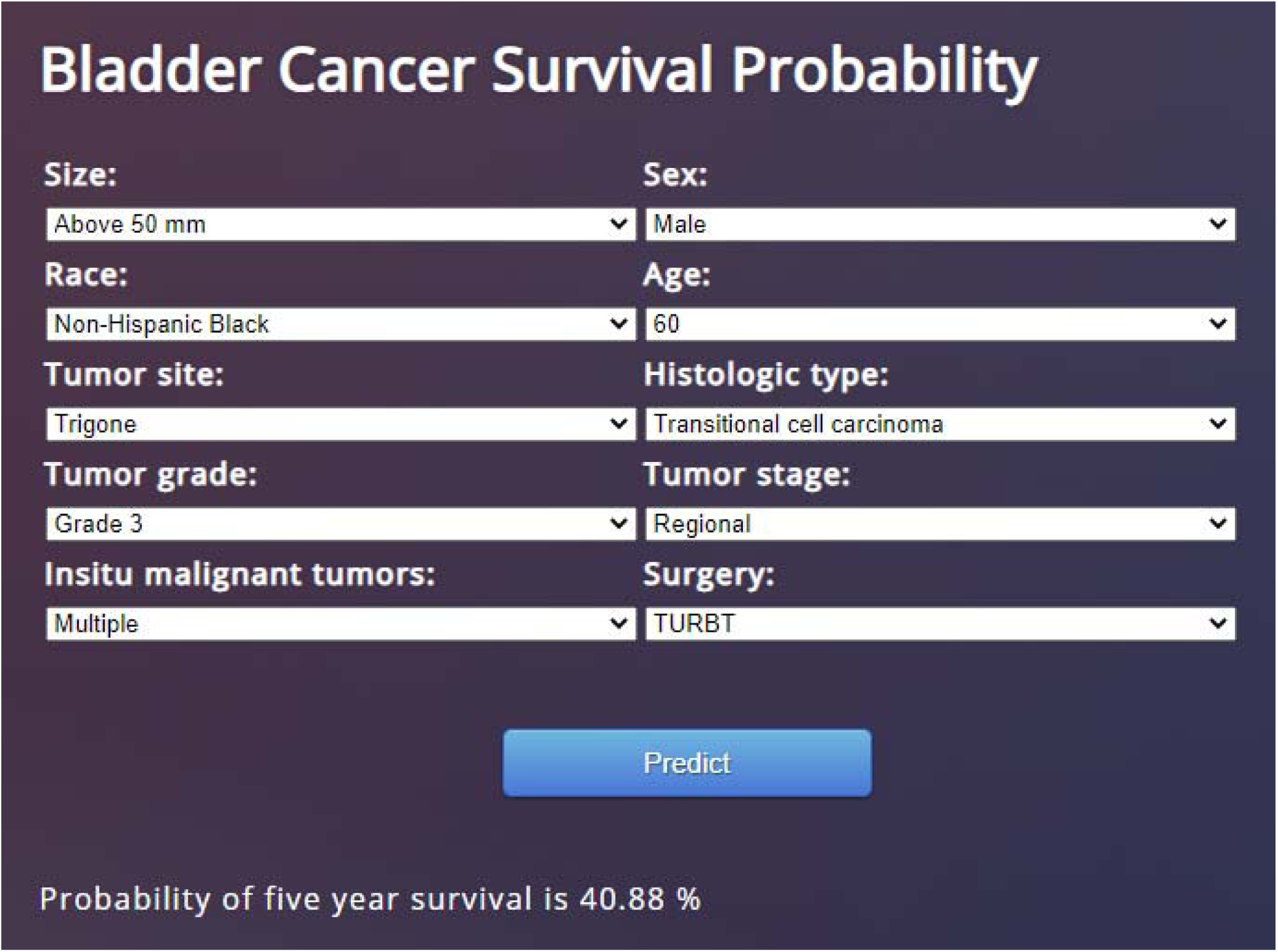

## Discussion

Despite advancements in early diagnosis and therapeutics, bladder cancer contributes to cancer burden worldwide, more so in high income countries.^29^ There are wider variations in the survival rates for bladder cancers by site in the US. For instance, the five-year survival rate for in situ tumors is 95.8%, whereas only 4.6% for metastatic disease.^30^ Therefore, it is essential for patients and providers to understand the prognosis of the cases including survival probability for further clinical management. In this study, we utilized machine learning algorithms to predict five-year survival among bladder cancer patients. Gradient boosting performed the best among all our algorithms and was deployed as an online survival prediction application for bladder cancer named BlaCaSurv.

There are at present a few predictive web applications using machine learning on SEER database that have been developed to predict cancer survival rate from spinal chordoma, chondrosarcoma, colorectal cancer, and glioblastoma.^15,31–33^ To the best of our knowledge, this is the first machine learning based online survival prediction application for bladder cancer patients. While using the SEER data base to predict five-year survival, Karhade et al. applied machine learning algorithms to 265 spinal chordoma patients and Thio et al. applied machine learning algorithms to 1,554 chondrosarcoma patients.^15,32^ They applied several algorithms – decision trees, neural networks, support vector machines, and Bayes point machines. However, both the studies reported Bayes point machine to be the best performer and used it for their web applications. Al-Bahrani et al. utilized deep neural network to predict survival of colon cancer patients, reported an AUC of 0.87 and have developed an online prediction application.^31^Another study by Senders et al. deployed multiple statistical and machine learning algorithms. Bagged decision trees, boosted decision trees, survival accelerated failure time (AFT), multilayer perceptron, boosted decision trees and support vector machines are a few among them.^33^ Their online prediction tool was AFT algorithm. The AUC for spinal chordoma was 0.868, for chondrosarcoma it was 0.8 AUC and glioblastoma had an AUC of 0.7. A recent study of bladder cancer survival among SEER patients using artificial neural network obtained an AUC of 0.81.^5^ The best performing model in our study however had an AUC of 0.927, which is the highest for any bladder cancer study using machine learning on SEER database.

Our study has several potential limitations. First, SEER data being an administrative database has its own biases and limitations – such as incomplete patient-level data on cancer risk and treatment, and inaccuracies and incompleteness of the source registries.^34^ Second, the database does not collect information on key socio-demographic features such as geographic location, household education and economic status as well as co-morbidities. All these additional clinical and socio-demographic factors can influence survival in bladder cancer patients. Inclusion of these additional features may improve the accuracy and reliability of the model.^35^ Finally, the application is yet to be externally validated using datasets outside the experimental cohort. Therefore, we urge caution while using this application as a predictive guide for ascertaining survival for bladder cancer. Clinicians could balance the predictions from this application against their clinical experience, relevant bio-chemical and radiologic parameters.

Our survival prediction application is one of the first such tools for bladder cancer in the United States. We have developed the application by using the largest cancer database in the US and hope this would be more generalizable. However, this application could further be validated and possibly reoptimized with the help of heterogenous data from multiple cohorts and multiple practice settings. Although external validation is required, this survival prediction application could be used as a supplementary tool to inform clinical decision-making for bladder cancer patients.

## Data Availability

Paper uses SEER database

## Authors’ contributions statement

**Ashis Kumar Das:** Conceptualization, study design, data acquisition, analysis, and writing– original draft. **Shiba Mishra:** Conceptualization, data interpretation, and writing–review and editing. **Devi Kalyan Mishra:** Data interpretation and writing–review and editing. **Saji Saraswathy Gopalan:** Conceptualization, data interpretation, and writing–review and editing.

## Acknowledgements

We are grateful to the contributors of the Surveillance, Epidemiology, and End Results Program as well as to the National Cancer Institute for making this data publicly available.

